# The association between SARS-CoV-2 infections in English primary and secondary school children and staff, and infections in members of their household in the schoolyear 2020-2021: a self-controlled case-series analysis

**DOI:** 10.1101/2023.11.27.23299066

**Authors:** Elliot McClenaghan, Patrick Nguipdop-Djomo, Alexandra Lewin, Charlotte Warren-Gash, Sarah Cook, Punam Mangtani

## Abstract

**Background:** The role of children and staff in SARS-CoV-2 transmission outside and within households is still not fully understood when large numbers are in regular, frequent contact in schools.

**Methods:** We used the self-controlled case-series method during the alpha- and delta-dominant periods to explore the incidence of infection in periods around a household member infection, relative to periods without household infection, in a cohort of primary and secondary English school children and staff from November 2020 to July 2021.

**Results:** We found the relative incidence of infection in students and staff was highest in the 1-7 days following household infection, remaining high up to 14 days after, with risk also elevated in the 6-12 days before household infection. Younger students had a higher relative incidence following household infection, suggesting household transmission may play a more prominent role compared with older students. The relative incidence was also higher amongst students in the alpha variant dominant period.

**Conclusions:** This analysis suggests SARS-CoV2 infection in children, young people and staff at English schools were more likely to be associated with within-household transmission than from outside the household, but that a small increased risk of seeding from outside is observed.

**Key messages:**

- **Question**: With respect to incidence of SARS-CoV-2 infection before and after household member infection, is within-household transmission more likely than community transmission amongst school children and staff?
- **Findings**: In this self-controlled case-series analysis, the relative incidence of infection in students and staff was highest in the 1-7 days following household infection, remaining high up to 14 days after, with the highest relative incidence found in younger, primary school-aged children.
- **Importance**: Within-household transmission is more likely than from outside the household, but a small increased risk of seeding from outside the household is observed as well as variation by age and variant dominant period.

## BACKGROUND

The first cases of severe acute respiratory coronavirus 2 (SARS-CoV-2) emerged in December 2019 and subsequently spread worldwide.^1^ In response to the significant morbidity and mortality caused by coronavirus (COVID-19) disease, many governments rapidly implemented non-pharmaceutical interventions (NPI) to curb transmission of SARS-CoV-2, which often included school closures. In the United Kingdom (UK) schools were intermittently closed to most students between March 2020 and September 2021. These closures have been associated with ongoing negative consequences on students’ learning and wellbeing.^2,3^

Initial caution that schools might contribute to community transmission was partly informed by influenza outbreaks where children are key drivers of transmission. There was however, early on, conflicting evidence of the importance of SARS-CoV-2 transmission by children and young people (CYP) in both households and school settings. This uncertainty together with the higher proportion of asymptomatic infection in CYP compared to adults led to epidemiological studies with active ascertainment such as the English Schools Infection Survey (SIS).^4–8^

A key question is the importance of school CYP and staff infections in schools, seeding household infections and helping sustain or amplify community transmission. The presence of household co-infections was the strongest risk factor in a study of determinants of incident SARS-CoV-2 infection in the English SIS.^9^ The direction of transmission within households was however not possible to investigate.

We investigated within-household transmission of SARS-CoV-2 amongst English primary and secondary school children and school staff through the application of self-controlled case series (SCCS) method to linked England SIS, national surveillance, and immunization data. We estimated the relative incidence of SARS-CoV-2 infection in SIS participants in different time periods before and after infection in a household member and investigated variations by age, household size, dominant SARS-CoV-2 variant of concern, and school type.

## METHODS

### Study Design and Participants

We used the SCCS method, where the incidence of events in risk periods following exposure is estimated and compared to that in other periods in the same person during a specified observation time.^10^ Participants were students aged 4-18 years and school staff members in the SIS cohort with recorded laboratory-confirmed SARS-CoV-2 infection and a record that another individual in their household had a SARS-CoV-2 infection between Nov 2020 and July 2021. The event (outcome) of interest was SARS-CoV-2 infection in the SIS participant, and the main exposure was the first reported SARS-CoV-2 infection (other than the SIS participant) in the household.

### Data Sources

Students and staff with laboratory-confirmed SARS-CoV-2 infection were identified from SIS. SIS was a longitudinal study during which biological samples and questionnaire data was collected from students and staff in six panel surveys at half-termly intervals, targeting 150 randomly selected state-funded primary and secondary schools across 15 Local Authorities (LAs) in England during the 2020-2021 school year; schools were selected using multi-stage stratified random sampling respectively in 10 Las from the top quintile and 5 LA from the bottom four quintiles community incidence in September 2020.^11,12^ Biological analyses included RT-PCR testing of nasal swabs for SARS-CoV-2 infection in those present in school on the visit day.

Participants’ records were linked to the UK Health Security Agency (UKHSA) Second Generation Surveillance System (SGSS) which contains nationwide infectious disease laboratory reports, including SARS-CoV-2 PCR and antigen tests results and symptoms status at the time of sampling including those in the SIS. The Unique Property Reference Numbers (UPRN) – which uniquely identify residential addresses in England, was used to anonymously link participants’ household members to SGSS results. SGSS data was used to obtain fact and date of infections in SIS participants and their household members. SIS questionnaires provided socio-demographic information and household size. The linked National Immunisation Management System (NIMS) provided COVID-19 vaccination status (for SIS participants only), and linked ONS census data gave an Index of Multiple Deprivation (IMD) score, a small-area indicator of socio-economic deprivation.^13^

The study was approved by UKHSA Research Support and Governance Office (NR0237) and London School of Hygiene & Tropical Medicine Ethics Review Committee (ref:22657).

### Outcome & Exposures

The main outcome was laboratory-confirmed SARS-CoV-2 infection in SIS participants during the 2020/2021 school year. Re-infections were uncommon during the study period, when the alpha- and delta-virus variants dominated, estimated at 0.68% in children and adults.^14^ The exposure event was restricted to the first reported household infections other than the SIS participant, assuming that temporally clustered subsequent cases within the household would either result from a chain of transmission within the household or common exposure to another source external to the household. Subgroup analyses were conducted separately for the periods when the alpha (infection reported before 1 May 2021) and delta (infection reported after 31 May 2021) virus variants dominated.^15^

### Definition of Risk Periods

The exposure periods were defined relative to the date the positive SARS-CoV-2 test sample was collected for the household member. The observation period was divided into a baseline and ‘higher’ risk periods (see *Figure 1*). The overall ‘higher risk’ period spanned from 12 days before the household member infection to 14 days after; it was further subdivided in four risk windows, respectively (i) -12 to -6 (a six-day ‘pre-exposure’ risk period, when infection introduction into the household by a SIS participant could occur as well as diagnosis and reporting delays of a household infection); (ii) -5 to 0 (the length includes 2-3 days pre-symptomatic infectiousness of the first household infection other than a SIS participant and 1-2 days until the test in a SIS participant was taken); (iii) 1 to 7, and (iv) 8 to 14 days after the household member infection, the latter to account for 14 days of infectiousness following infection.^16–18^

**Figure 1:**
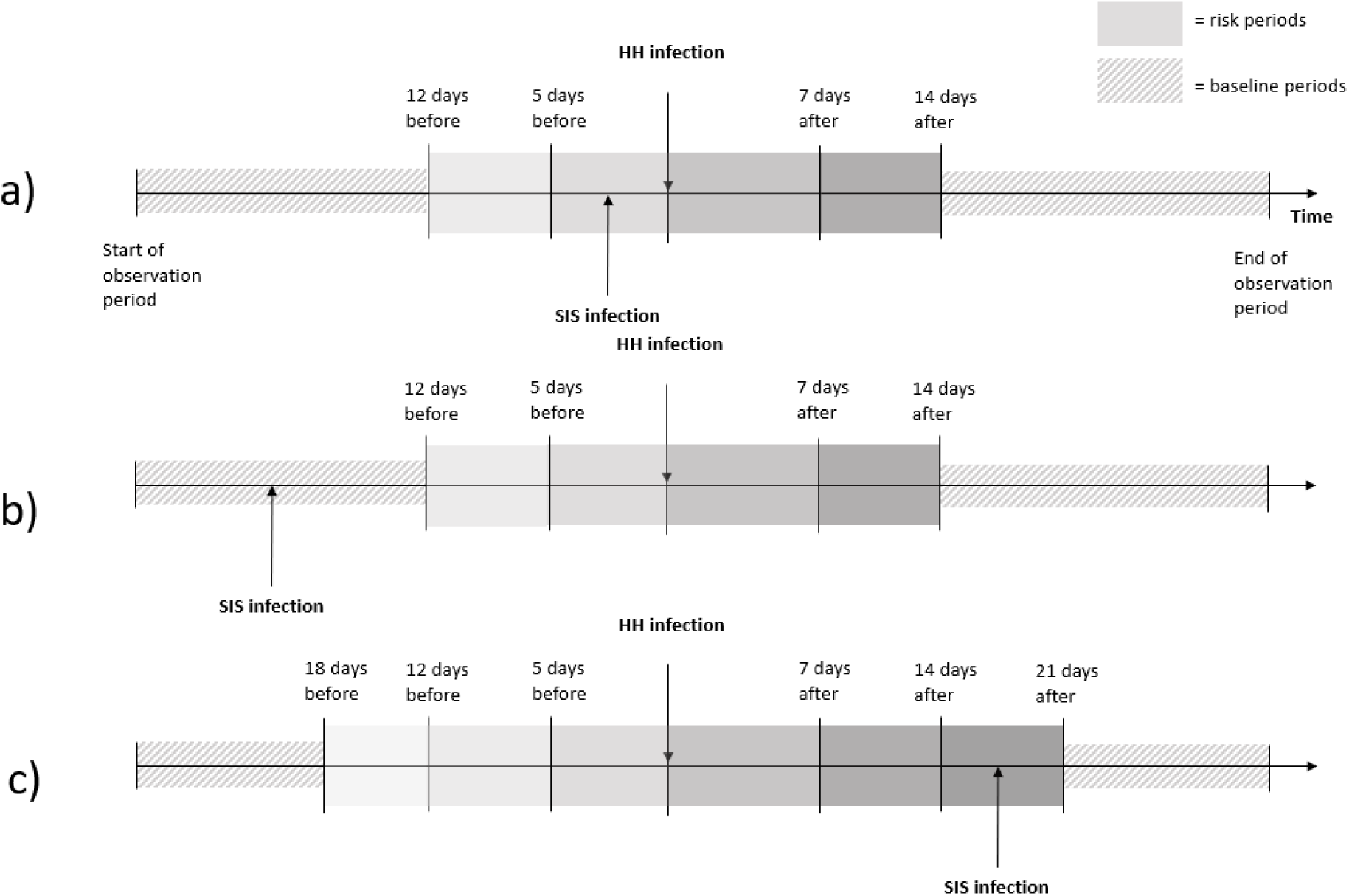
Study design for investigating the association between survey participant infection and household infection, showing the 26-days interval with four risk periods (a and b) and the 39-days interval with six risk periods (c). Abbreviations: HH, household; SIS, school infection survey; SCCS, self-controlled case-series. These diagrams are not to scale, and placement of the household (HH) infection (exposure) is for illustrative purposes - exposures are transient and can occur anywhere in the observation period (2 November 2020 to 6 July 2021). Each diagram represents the observation period of one SIS participant. The main analysis **(a)** and **(b)** models 26-days interval with four exposure risk periods spanning (i) 6-12 days before HH infection, (ii) 0-5 days before HH infection, (iii) 1-7 days following HH infection and (iv) 8-14 days following HH infection. Exposure risk estimates are produced using the baseline periods (all parts of the observation period outside the risk period) as the reference group. In **(a)** the SIS infection (outcome event) occurs within the 0-5 days preceding HH infection. In **(b)** the SIS infection occurs outside the risk periods in the baseline period. In contrast, the sensitivity analysis **(c)** models a 39-days interval with six risk periods defined, including an additional six (13-18) days before HH infection, and seven (15 to 21) days after the HH infection. In all examples, the observation period following event is modelled using a pseudo-loglikelihood as per the SCCS counterfactual method extension.

To check the robustness of our exposure risk period definition, we explored a longer 39-days interval, including an additional six days (-18 to -13) before, and seven days (15 to 21) after the exposure events.

### Statistical Analysis

SCCS statistical models use a modified Poisson cohort design, conditioning on the occurrence of outcome events (SIS-1 participant infection), with each individual acting as their own control during the different exposure and baseline time periods. The outcome incidence in SIS participants within the defined risk windows is compared to that in the baseline period by estimating incidence rate ratios (IRR). We implemented an extension of the standard SCCS for censored post-event exposures^19^, to address the change in risk of (re)infection following infection (see *Supplementary figure S4*). Analyses were controlled for time-varying confounding on calendar time and vaccination status in staff (CYP were not yet eligible for vaccination). The SCCS model automatically controls for all observed and unobserved time-invariant confounders.

We divided the observation periods into three calendar time intervals, respectively 2 November 2020 to 1 January 2021 (schools reopening following the pandemic 1^st^ wave, subsequent school-closures and emergence of the alpha variant), 2 January 2021 to 1 March 2021 (2^nd^ national lockdown and phased roll-out of adult vaccination), and 2 March 2021 to 6 July 2021 (schools re-opening, introduction of mass testing). Individual vaccination status in staff, was adjusted for by dividing individual observation periods into unvaccinated and vaccinated intervals (>14 days following first vaccine dose).

We estimated the IRR separately in students and in staff, comparing the 4 risk windows (*Figure 1*) to the baseline period. We also investigated potential differences in IRRs by variant-dominant period, first household member infection age group and symptomatic status, household size, participant sex and school type (primary or secondary) by examining effect modification by these factors. A subgroup sensitivity analysis was restricted to study participants with positive PCR nasal swab only during SIS panel surveys.

All data management, linkage and statistical analysis were done within the ONS Secure Research Service (SRS) trusted research environment and performed using Stata version 17.^20^

## RESULTS

### Cohort characteristics

Overall, 5.8% (859/14,842) students and 10.4% (808/7743) staff who participated in the Schools Infection Survey had a lab-confirmed SARS-CoV-2 infection recorded in SGSS between 02 November 2020 and 06 July 2021. Of these, 957 people, respectively 439 (54.3%) staff and 518 (60.3%) students who had at least one household member infection recorded in SGSS, were included in our study. The study participants’ characteristics are presented in *Table 1*. About half of students were male, 39% were aged under 12 years, and 81% from White ethnicity; 40% lived in households with five or more members. Staff participants were mostly female (80%) and from White ethnicity (88%), with 57% aged ≥40 years and 70% working in secondary schools; 92% had received at least one COVID-19 vaccination by the end of the observation period.

**Table 1:** Descriptive characteristics of school infection survey participants testing positive for SARS-CoV-2 and experiencing a household infection in the observation period, Nov 2020 to July 2021 (N=957).

| Variable | Staff | Students |
| --- | --- | --- |
| Positive for SARS-CoV-2; N | 439 | 518 |
| Multiple HH infections; n (%) | 190 (43.3) | 328 (63.3) |
| No. HH infections; n (%) |  |  |
| 1-2 | 362 (82.5) | 370 (71.4) |
| 3-4 | 70 (15.9) | 138 (26.6) |
| 5-8 | 7 (1.6) | 10 (1.9) |
| Male sex; n (%) | 89 (20.3) | 240 (46.3) |
| Age (years)<sup>a</sup>; |  |  |
| <12 | 0 (0.0) | 203 (39.2) |
| 12-19 | 3 (0.7) | 315 (60.8) |
| 20-39 | 187 (42.6) | 0 (0.0) |
| 40+ | 249 (56.7) | 0 (0.0) |
| IMD Quintiles; n (%) |  |  |
| 1 (least deprived) | 76 (17.3) | 136 (26.3) |
| 2 | 81 (18.5) | 120 (23.2) |
| 3 | 84 (19.1) | 97 (18.7) |
| 4 | 109 (24.8) | 78 (15.1) |
| 5 (most deprived) | 84 (19.1) | 86 (16.6) |
| Ethnicity; n (%) |  |  |
| Asian | 36 (8.2) | 56 (10.8) |
| Black | 3 (0.7) | 10 (1.9) |
| Mixed | 8 (1.8) | 24 (4.6) |
| Other | * (-) | * (-) |
| White | 387 (88.2) | 420 (81.1) |
| Unknown | * (-) | * (-) |
| School type; n (%) |  |  |
| Primary | 131 (29.8) | 116 (22.4) |
| Secondary | 308 (70.2) | 402 (77.6) |
| SARS-CoV-2 variant dominant periods; n (%) |  |  |
| Before 01 May 2021 (Alpha) | 374 (85.2) | 348 (67.2) |
| May 2021 | 9 (2.1) | 19 (3.7) |
| After 31 May 2021 (Delta) | 56 (12.8) | 151 (29.2) |
| Term infection reported; n (%) |  |  |
| Autumn term<sup>b</sup> | 237 (54.0) | 173 (33.4) |
| Spring term<sup>c</sup> | 136 (31.0) | 174 (33.6) |
| Summer term<sup>d</sup> | 66 (15.0) | 171 (33.0) |
| Vaccinated in the observation period<sup>e</sup>; n (%) | 404 (92.0) | 26 (5.0) |
| Vaccinated before SARS-CoV-2 infection in participant;<br>n (%) | 57 (13.0) | * (-) |
| HH size; n (%) |  |  |
| 1-2 | 100 (22.8) | 31 (6.0) |
| 3-4 | 255 (58.1) | 280 (54.1) |
| 5+ | 83 (18.9) | 206 (39.8) |
| Characteristics of 1st HH infection |  |  |
| Age <18 years | 121 (27.6) | 121 (23.4) |
| Age 18+ years | 318 (72.4) | 397 (76.6) |
| Asymptomatic | 91 (20.7) | 135 (26.1) |
| Symptomatic | 330 (75.2) | 353 (68.1) |
| Unknown symptom status | 18 (4.1) | 30 (5.8) |

Abbreviations: HH, household; IMD, Index of Multiple Deprivation; SARS-CoV-2, severe acute respiratory coronavirus 2.
<sup>a</sup>age at enrollment into SIS-1 (31 August 2020) <sup>b</sup>3 November 2020 – 2 January 2021 <sup>c</sup>3 January 2021 – 17 April 2021 <sup>d</sup>18
April – 6 July 2021 <sup>e</sup>first dose received in the observation period. Vaccine offered to CYP only to those at high risk then
hence the small numbers. \*Suppressed for data protection purposes.

### Association between household member and participant SARS-CoV-2 infections

After adjusting for calendar time, and vaccination status in staff, the IRR of infection in all participants was higher in all risk windows (12 days before to 14 days after the first household infection), compared to the baseline periods (*Figure 2*). In both staff and students, the highest IRR was observed in the 1-7 days after the household member infection (staff IRR 93.95 (95%CI 65.59 to 134.59); students IRR 150.32 (95%CI 104.59 to 216.03)). A small increased relative incidence was seen in days -12 to -6 in students, and days -12 to -6 and 8 to 14 in staff but much lower than in the other periods (*Figures 3-4*).

**Figure 2:**
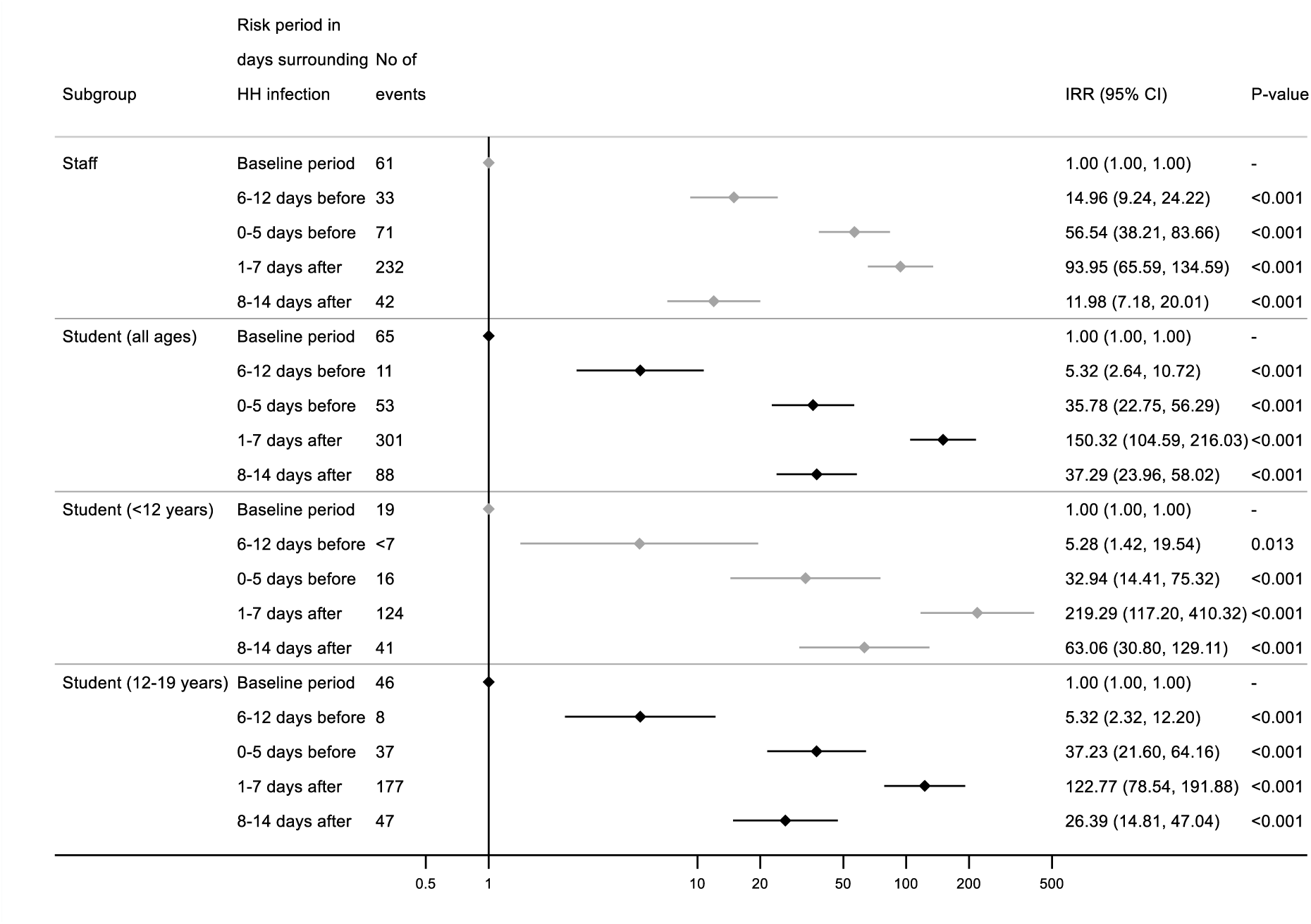
Incidence rate ratios (IRR) for school infection survey participant infection in relation to a participant household infection, stratified by staff, students, and student age groups, Nov 2020 to July 2021. Abbreviations: CI, confidence interval; HH, household; IRR, incidence rate ratio; SIS, school infection survey. No of events refers to number of SIS participant infections occurring across the baseline and risk periods. IRRs are adjusted for calendar time (all subgroups) and vaccination status (staff only). Wald test two-sided p-values.

**Figure 3:**
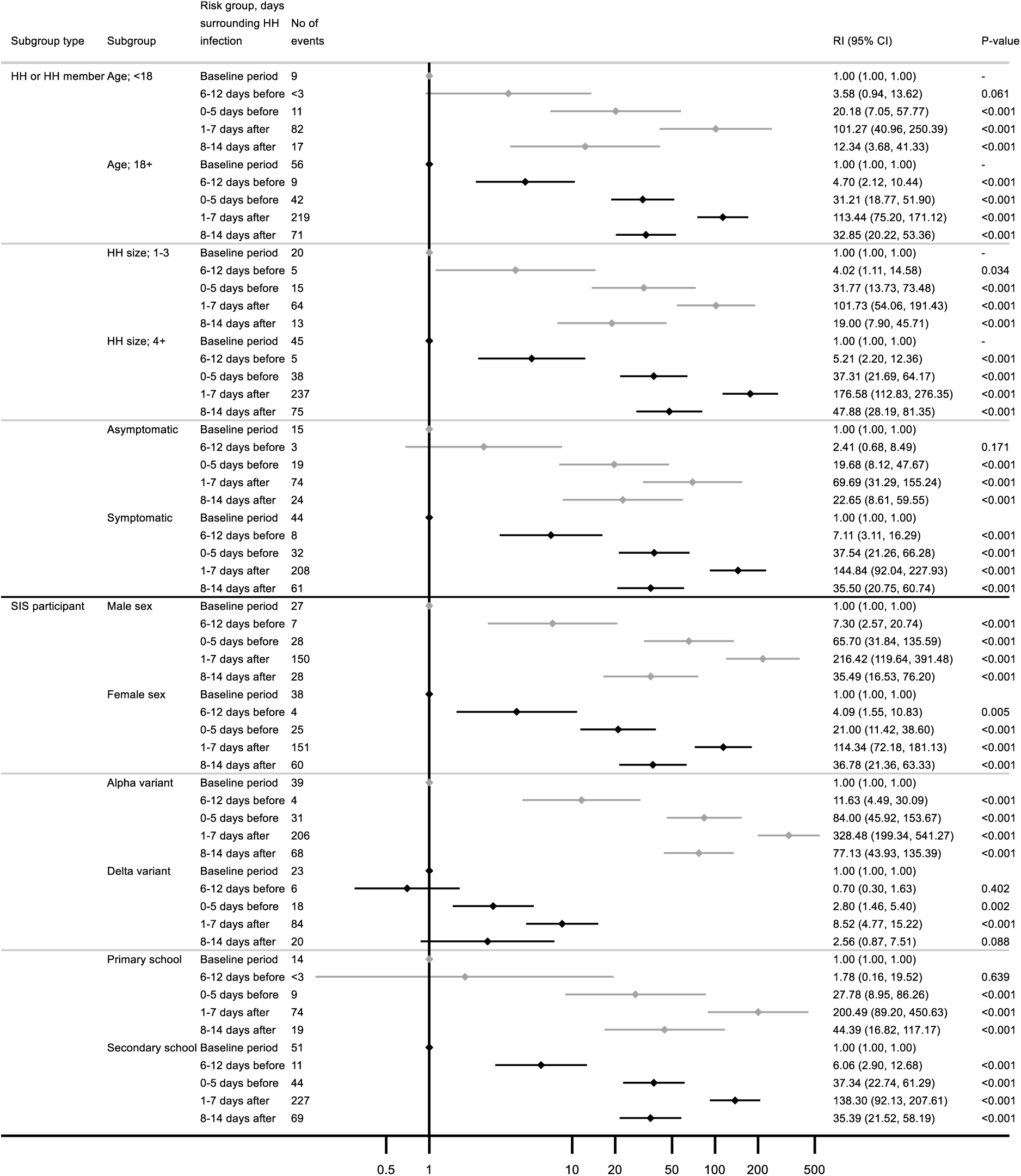
Incidence rate ratios (IRR) for student (N=518) school infection survey participant infection in relation to household infection by household member and participant characteristics, Nov 2020 to July 2021. Abbreviations: CI, confidence interval; HH, household; IRR, incidence rate ratio; SIS, school infection survey. For the variant-dominant period subgroups, calendar time is not adjusted for as a time varying confounder in the model. No of events refers to number of SIS infections occurring across the baseline and risk periods. IRRs are adjusted for calendar time (all subgroups except variants). Wald test two-sided p-values.

**Figure 4:**
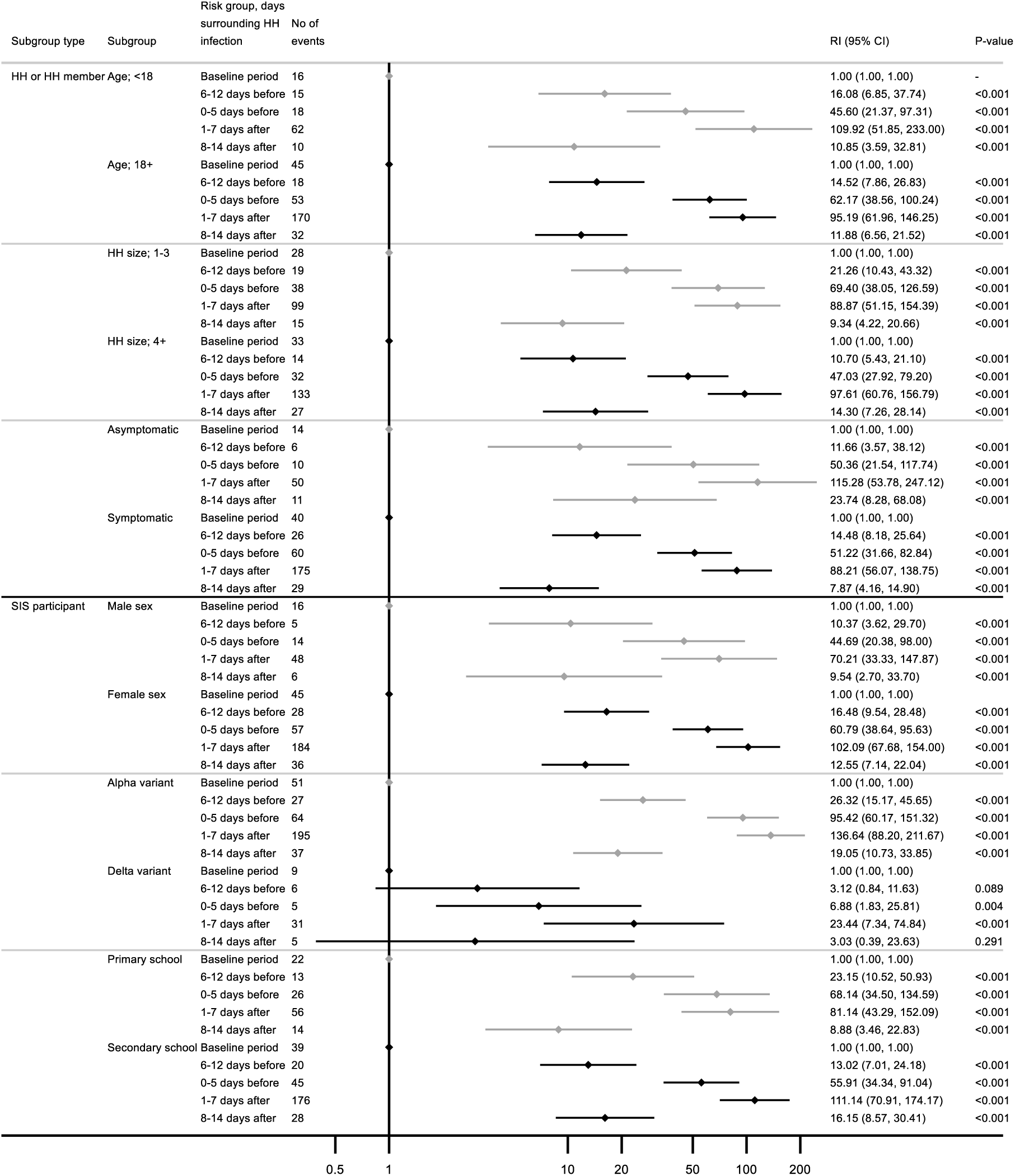
Incidence rate ratios (IRR) for staff (N=439) school infection survey participant infection in relation to household infection across household member characteristics and participant characteristics, Nov 2020 to July 2021. Abbreviations: CI, confidence interval; HH, household; IRR, incidence rate ratio; SIS, school infection survey. For the variant-dominant period subgroups, calendar time is not adjusted for as a time varying confounder in the model. No of events refers to number of SIS infections occurring across the baseline and risk periods. IRRs are adjusted for calendar time (all subgroups except variants) and receipt of first dose vaccination. Wald test two-sided p-values.

We further stratified the analysis in students by age group, respectively under 12 years (typically primary school) and 12 to 19 years (typically secondary school) (*Figure 2).* The overall relative incidence pattern was the same in both groups and similar to all students; however, the higher relative incidence observed 1-7 days and 8-14 days after the household member infection were nearly twice as high in younger (<12 years) students than 12-19 years students, suggesting higher rate of infection in younger children following a household case, than older students.

Comparing IRRs using a wider (39 days) exposure window to our main analysis’ 26 days window, showed much lower relative risks in the additional exposure days supporting our exposure window definition (details in *Supplementary table S1*). Full results for the sensitivity analyses for the wider exposure period and the restricted subgroup with PCR-confirmed infection during the SIS panel surveys can be found in *Supplementary table S1* and *Supplementary table S3*, respectively.

We also investigated differences in the association between household member and participant’s infection by various participant (sex, age, household size (≤3 versus >3 members), school type (primary vs secondary), dominant SARS-CoV-2 variant (Alpha versus delta)) and household member (age (<18 vs ≥18 years) and symptomatic status) characteristics (*Figures 3-4 and Supplementary table S2).* In staff, the pattern and magnitude of associations were broadly comparable across all characteristics, except the dominant virus variant periods for which IRRs were much higher during the alpha-dominant period (eg. day 1 to 7 IRR = 136.6 (95%CI 88.2 to 211.7)) than the delta-dominant period (eg. day 1 to 7 IRR = 23.4 (95%CI 7.3 to 74.8). The findings were similar in students.

## DISCUSSION

This study used a novel approach to investigate SARS-CoV-2 transmission in households with CYP and staff attending school for some of the SIS study period using a case-only design. The method was applied to linked data from a large national survey of school students and staff in England during the alpha- and delta-variant dominant periods in the 2020/2021 school year. In both students and staff, we found higher incidence of infection in the 1 to 7 days and 8 to 14 days following infection of a household member, compared to periods when there was no infection in the household, with the relative incidence highest in students aged under 12 years, and higher during the alpha-dominant period. We note the increased relative incidence, albeit consistently lowest, in the 6 to 12 days before the household case in all participants; this may reflect situations where the household member acquired the infection from the SIS cohort study participant or another source, compatible with infections in schools seeding into households and a chain of transmission in the household though to a lesser extent than within household transmission.

Schools were closed because of concerns that educational settings could sustain or amplify the transmission of SARS-CoV-2 in the community, further compounded by the recognition through surveillance that infections with little to no symptoms were frequent, especially in younger populations, yet they could be infectious.^4,21–23^ Respiratory infections that are preferentially aerosol transmitted require close contact. Consistent with this, household transmission was noted to be more important for SARS-CoV-2 than school transmission in a meta-analysis.^22^

The finding in our study that the relative incidence of infection in our participants, both students and staff, was consistently highest in the week following the first case in their household, and much lower in the 12 days before, is consistent with the hypothesis that school participants acquired infection predominantly from within their household, than out of the household. Several studies have noted the high within-household clustering of SARS-CoV-2 infections, including during ‘lockdown’ periods when community mixing was severely restricted. Schools remained one of the few settings where high frequency of close contacts between large numbers of people was possible.^9,24,25^ In the SIS study period, there were several times when schools were closed leading to increased mixing at home, which may have facilitated household transmission.

The results are also consistent with SARS-CoV-2 alpha and delta variants’ generation time.^17^ Previous studies have shown SARS-CoV-2 to be infectious up to 7-10 days following reporting of symptoms, and that infectiousness rarely continues more than 10 days.^26,27^ This is reflected in our findings with IRRs attenuated towards the null in the 8-14 days following household infection, but the evidence for an increased IRR in this period compared with baseline is still pronounced, and may be consistent with some delay from testing and reporting.^28,29^

The highest relative incidence in the week following the household case was inversely correlated to participant’s age, highest in younger students, and decreasing in older students and school staff. This result is compatible with older individuals more able to modulate their behaviour to reduce the risk of transmission when there is a diagnosed infectious person in their household, than younger people, and also the fact younger people require care, thus more contact with other members of the household, than older subjects. Other studies have noted that children under 10-12 years have increased susceptibility to transmission.^23^

The relative incidences in the periods after household infection were consistently lower during the delta verses the alpha variant dominant periods, across all participants and age groups. Estimates of transmissibility based on household secondary attack rates (SARs) were previously noted to vary during the alpha- and delta-dominant waves.^24,25,30^The higher infectiousness of delta compared to alpha may have increased community transmission compared with household transmission.^17^ Moreover, the relaxation of restrictions to social mixing in England around the time delta was spreading in the Spring and beginning of Summer 2021, could have led to greater opportunities for acquisition of infection out of the household leading to lower apparent within-household relative incidence.

Overall, our analyses support the idea that during the 2020-2021 school year in England, primary and secondary schools’ students and staff were more likely to have acquired infection from within their household than out. A limitation is that we did not have the data to directly measure the incidence of infection in the household following infection in a school participant. Incomplete household testing and reporting after a first reported case was possible. We instead approximated the IRR in school participants in the 6 to 12 days before the first household case. Alternative study designs have also struggled to measure household attack rates, with for example household contact studies being labor-intensive and expensive, with relatively low yield and power.^6,31^

Another potential limitation in our analysis is that although testing behavior was broadly similar throughout the observation period, it may have varied due to local epidemic trends and government recommendations. In an analysis conducted for the UK government’s Events Research Program where risk of SARS-CoV-2 was assessed following attendance of cultural events using SCCS, this potential testing variability was controlled for by dividing the rate ratio for positive tests by the rate ratio for negative tests over the same period and producing adjusted estimates.^32^ Lack of complete negative testing data precluded similar adjustments in our study. We did split our observation period to adjust for calendar time, but there is still potential for residual time-varying confounding given the rapid changes to epidemic conditions during the study period.

Despite its limitations, the case-only design used in this study had several strengths, including our ability to use readily available surveillance data, and the design’s intrinsic adjustment for both measured and unmeasured between-person fixed confounders, such as age, sex, and risk behavior. While not perfect, this design allowed to use of routinely collected surveillance data to capture household and individual infection, where there may be less capture of multiple infections.

This the case-only method (SCCS) can be a valuable method for similar analyses of large datasets of test results and surveillance surveys where detailed parameters on transmission dynamics may not be available, to generate early insights into the relative incidence of household compared to community transmission of infectious outbreaks such as SARS-CoV-2. Further research could usefully include similar analyses during the Omicron and future variant waves, incorporation of participant and household member re-infections and genomic data, as well as further exploration of household size and assessment of impact of interventions to mitigate transmission in schools as well as other frontline workers. This study provides evidence for a greater magnitude of an association of SARS-CoV-2 infection with household co-infection in CYP than by attending English schools with the risk in households varying by age and variant-dominant periods.

## Data Availability

De-identified study data are available for access by accredited researcher in the ONS Secure Research Service (SRS) for accredited research purposes under part 5, chapter 5 of the Digital Economy Act 2017. For further information about accreditation, contact or visit the SRS website.

## Author Notes

## Acknowledgements

This study was funded by the UK Department of Health and Social Care. We would like to thank the schools, head teachers, staff, families and children who took part in the SIS study. We are grateful to the team at IQVIA and to the SIS Engagement Officers for working tirelessly in communicating with and supporting the schools. For the purpose of Open Access, the author has applied a CC BY public copyright licence to any Author Accepted Manuscript (AAM) version arising from this submission.

## Disclaimer

The data used in this report is independent research funded by the Department of Health and Social Care (COVID 19-NTP 2.0, School Infection Study). The views expressed in this publication are those of the author(s) and not necessarily those of the NHS, or the Department of Health and Social Care.

## Authors contribution statement

PND and PM conceived the study design and methodology, and were involved in acquisition of the data with SC. EM performed the analysis, with support from PND, PM, SC, CWG and AL. EM, PND, and PM, drafted the manuscript. All authors gave critical revision of the manuscript for important intellectual content and contributed to reviewing and editing of the manuscripts. All authors had access to the data. EM and PND had final responsibility to submit for publication.

## Abbreviations

COVID-19: coronavirus-19;
CYP: children and young people;
IMD: Index of Multiple Deprivation
IRR: incidence rate ratio;
LA: Local Authority;
NIMS: National Immunisation Management System;
ONS: Office for National Statistics;
PCR: polymerase chain reaction;
RT-PCR: reverse transcription polymerase chain reaction;
SARS-CoV-2: severe acute respiratory coronavirus 2;
SCCS: self-controlled case-series;
SGSS: Second Generation Surveillance System;
SIS: school infection survey;
SRS: Secure Research Service;
UK: United Kingdom;
UKHSA: United Kingdom Health Security Agency;
WHO: World Health Organization

## Data and Statistical analysis code availability statement

The statistical analysis code is publicly available at github.com/ElliotMcC/SIS_SCCS.

## Source, licence copyrights and credits

This work contains statistical data from ONS which is Crown Copyright. The use of the ONS statistical data in this work does not imply the endorsement of the ONS in relation to the interpretation or analysis of the statistical data. This work uses research datasets which may not exactly reproduce National Statistics aggregates.

## Ethics approval

The Public Health England (now part of the UK Health Security Agency) Research Support and Governance Office (NR0237) and the London School of Hygiene & Tropical Medicine Ethics Review Committee (ref: 22657).

## References

1. Guan W-J, Ni Z-Y, Hu Y, et al. Clinical Characteristics of Coronavirus Disease 2019 in China. New England Journal of Medicine. 2020;382(18):1708–1720. doi:10.1056/nejmoa2002032

2. Viner R, Russell S, Saulle R, et al. School Closures During Social Lockdown and Mental Health, Health Behaviors, and Well-being Among Children and Adolescents During the First COVID-19 Wave. JAMA Pediatrics. 2022;176(4):400. doi:10.1001/jamapediatrics.2021.5840

3. Sundaram N, Abramsky T, Oswald WE, et al. Implementation of <scp>COVID</scp>-19 Preventive Measures and Staff Well-Being in a Sample of English Schools 2020-2021. Journal of School Health. 2023;93(4):266–278. doi:10.1111/josh.13264

4. Flasche S, Edmunds WJ. The role of schools and school-aged children in SARS-CoV-2 transmission. The Lancet Infectious Diseases. 2021;21(3):298–299. doi:10.1016/s1473-3099(20)30927-0

5. Forbes H, Morton CE, Bacon S, et al. Association between living with children and outcomes from covid-19: OpenSAFELY cohort study of 12 million adults in England. BMJ. 2021:n628. doi:10.1136/bmj.n628

6. House T, Riley H, Pellis L, et al. Inferring risks of coronavirus transmission from community household data. Statistical Methods in Medical Research. 2022;31(9):1738–1756. doi:10.1177/09622802211055853

7. Af Geijerstam A, Mehlig K, Hunsberger M, Åberg M, Lissner L. Children in the household and risk of severe COVID-19 during the first three waves of the pandemic: a prospective registry-based cohort study of 1.5 million Swedish men. BMJ Open. 2022;12(8):e063640. doi:10.1136/bmjopen-2022-063640

8. Flook M, Jackson C, Vasileiou E, et al. Informing the public health response to COVID-19: a systematic review of risk factors for disease, severity, and mortality. BMC Infectious Diseases. 2021/04/12 2021;21(1):342. doi:10.1186/s12879-021-05992-1

9. Nguipdop-Djomo P, Oswald WE, Halliday KE, et al. Risk factors for SARS-CoV-2 infection in primary and secondary school students and staff in England in the 2020/2021 school year: a longitudinal study. International Journal of Infectious Diseases. 2023;128:230–243. doi:10.1016/j.ijid.2022.12.030

10. Petersen I, Douglas I, Whitaker H. Self controlled case series methods: an alternative to standard epidemiological study designs. BMJ. 2016:i4515. doi:10.1136/bmj.i4515

11. al He. The COVID-19 Schools Infection Survey in England: Protocol and participation profile for a prospective, observational cohort study. 2022;doi:10.2196/34075

12. Hargreaves JR, Langan SM, Oswald WE, et al. Epidemiology of SARS-CoV-2 infection among staff and students in a cohort of English primary and secondary schools during 2020–2021. The Lancet Regional Health - Europe. 2022;21:100471. doi:10.1016/j.lanepe.2022.100471

13. David McLennan SN, Michael Noble, Emma Plunkett, Gemma Wright and Nils Gutacker. English Indices of Deprivation 2019: technical report. 2019. 2019. https://assets.publishing.service.gov.uk/government/uploads/system/uploads/attachment_data/file/833951/IoD2019_Technical_Report.pdf

14. Mensah AA, Campbell H, Stowe J, et al. Risk of SARS-CoV-2 reinfections in children: a prospective national surveillance study between January, 2020, and July, 2021, in England. The Lancet Child & Adolescent Health. 2022;6(6):384–392. doi:10.1016/s2352-4642(22)00059-1

15. England PH. SARS-CoV-2 variants of concern and variants under investigation in England Technical briefing 23. 2021. 17 September 2021. Accessed 6th June 2023. https://assets.publishing.service.gov.uk/government/uploads/system/uploads/attachment_data/file/1018547/Technical_Briefing_23_21_09_16.pdf

16. Casey M, Griffin J, McAloon CG, et al. Pre-symptomatic transmission of SARS-CoV-2 infection: a secondary analysis using published data. Cold Spring Harbor Laboratory; 2020.

17. Hart WS, Miller E, Andrews NJ, et al. Generation time of the alpha and delta SARS-CoV-2 variants: an epidemiological analysis. The Lancet Infectious Diseases. 2022;22(5):603–610. doi:10.1016/s1473-3099(22)00001-9

18. Byrne AW, McEvoy D, Collins AB, et al. Inferred duration of infectious period of SARS-CoV-2: rapid scoping review and analysis of available evidence for asymptomatic and symptomatic COVID-19 cases. BMJ Open. 2020;10(8):e039856. doi:10.1136/bmjopen-2020-039856

19. Farrington CP, Whitaker HJ, Hocine MN. Case series analysis for censored, perturbed, or curtailed post-event exposures. Biostatistics. 2008;10(1):3–16. doi:10.1093/biostatistics/kxn013

20. Stata Statistical Software: Release 17. StataCorp LLC; 2021.

21. Cordery R, Reeves L, Zhou J, et al. Transmission of SARS-CoV-2 by children to contacts in schools and households: a prospective cohort and environmental sampling study in London. The Lancet Microbe. 2022;3(11):e814–e823. doi:10.1016/s2666-5247(22)00124-0

22. Viner R, Waddington C, Mytton O, et al. Transmission of SARS-CoV-2 by children and young people in households and schools: A meta-analysis of population-based and contact-tracing studies. Journal of Infection. 2022;84(3):361–382. doi:10.1016/j.jinf.2021.12.026

23. Viner RM, Mytton OT, Bonell C, et al. Susceptibility to SARS-CoV-2 Infection Among Children and Adolescents Compared With Adults. JAMA Pediatrics. 2021;175(2):143. doi:10.1001/jamapediatrics.2020.4573

24. Toth DJA, Beams AB, Keegan LT, et al. High variability in transmission of SARS-CoV-2 within households and implications for control. PLOS ONE. 2021;16(11):e0259097. doi:10.1371/journal.pone.0259097

25. Madewell ZJ, Yang Y, Longini IM, Halloran ME, Dean NE. Household Transmission of SARS-CoV-2. JAMA Network Open. 2020;3(12):e2031756. doi:10.1001/jamanetworkopen.2020.31756

26. Adam D. How long is COVID infectious? What scientists know so far. News. Nature. 26 July 2022 2022;608:16–17. 10.1038/d41586-022-02026-x

27. Townsley H, Carr EJ, Russell TW, et al. Non-hospitalised, vaccinated adults with COVID-19 caused by Omicron BA.1 and BA.2 present with changing symptom profiles compared to those with Delta despite similar viral kinetics. Cold Spring Harbor Laboratory; 2022.

28. Tu H, Hu K, Zhang M, Zhuang Y, Song T. Effectiveness of 14 day quarantine strategy: Chinese experience of prevention and control. BMJ. 2021:e066121. doi:10.1136/bmj-2021-066121

29. Mahase E. Covid-19: Is it safe to reduce the self-isolation period? BMJ. 2021:n3164. doi:10.1136/bmj.n3164

30. Fung HF, Martinez L, Alarid-Escudero F, et al. The Household Secondary Attack Rate of Severe Acute Respiratory Syndrome Coronavirus 2 (SARS-CoV-2): A Rapid Review. Clinical Infectious Diseases. 2020;73(Supplement_2):S138–S145. doi:10.1093/cid/ciaa1558

31. Derqui N, Koycheva A, Zhou J, et al. Risk factors and vectors for SARS-CoV-2 household transmission: a prospective, longitudinal cohort study. The Lancet Microbe. 2023;4(6):e397–e408. doi:10.1016/s2666-5247(23)00069-1

32. Department for Digital C, Media & Sport, GOV.UK. Science Note - A self-controlled case series study to measure the risk of SARS-CoV-2 infection associated with attendance at an Events Research Programme event. gov.uk. Accessed 9 November 2022, 2022. https://www.gov.uk/government/publications/events-research-programme-phase-ii-and-iii-findings/science-note-a-self-controlled-case-series-study-to-measure-the-risk-of-sars-cov-2-infection-associated-with-attendance-at-an-events-research-progra

