## Supplementary materials for "The association between SARS-CoV-2 infections in English primary and secondary school children and staff, and infections in members of their household in the schoolyear 2020-2021: a self-controlled case-series analysis"

### Article Title:

### Table of contents:

*Supplementary figure S1*: Histogram showing the distribution of laboratory-confirmed SARS-CoV-2 infections amongst student school infection survey participants in the observation period, Nov 2020 to July 2021 (N=518).

*Supplementary figure S2*: Histogram showing the distribution of laboratory-confirmed SARS-CoV-2 infections amongst staff school infection survey participants in the observation period, Nov 2020 to July 2021 (N=439).

*Supplementary figure S3*: Histograms showing the number of days between participant household infections and school infection survey participant infection for staff and students, Nov 2020 to July 2021.

*Supplementary figure S4*: Detail and purpose of the SCCS counterfactual extension used in the main analysis.

*Supplementary table S1*: Incidence rate ratios (IRR) school infection survey participant infection in relation to a participant household infection, stratified by staff and students, including additional risk periods 13-18 days pre-household infection and 15-21 following household infection, Nov 2020 to July 2021.

*Supplementary table S2*: Incidence rate ratios (IRR) school infection survey participant infection in relation to participant household infection, stratified by staff and students and by potential effect modifiers of interest, using two risk groups amongst subgroups (in the four-risk-period main analysis, data sparsity did not allow examination of subgroups for age in staff), Nov 2020 to July 2021.

*Supplementary table S3*: Incidence rate ratios (IRR) school infection survey participant infection in relation to a participant household infection, stratified by staff, students, and student age groups, restricted to only PCR-confirmed SARS-CoV-2 infections recorded in the SIS panel surveys, Nov 2020 to July 2021.

*Supplementary table S4*: Incidence rate ratios (IRR) school infection survey participant infection in relation to a participant household infection, stratified by staff, students, and student age groups, using the standard self-controlled case-series analysis (ignoring the limiting assumption that observation period should not depend on events), Nov 2020 to July 2021.

*Supplementary figure S1*: Histogram showing the distribution of laboratory-confirmed SARS-CoV-2 infections amongst student school infection survey participants in the observation period, Nov 2020 to July 2021 (N=518).

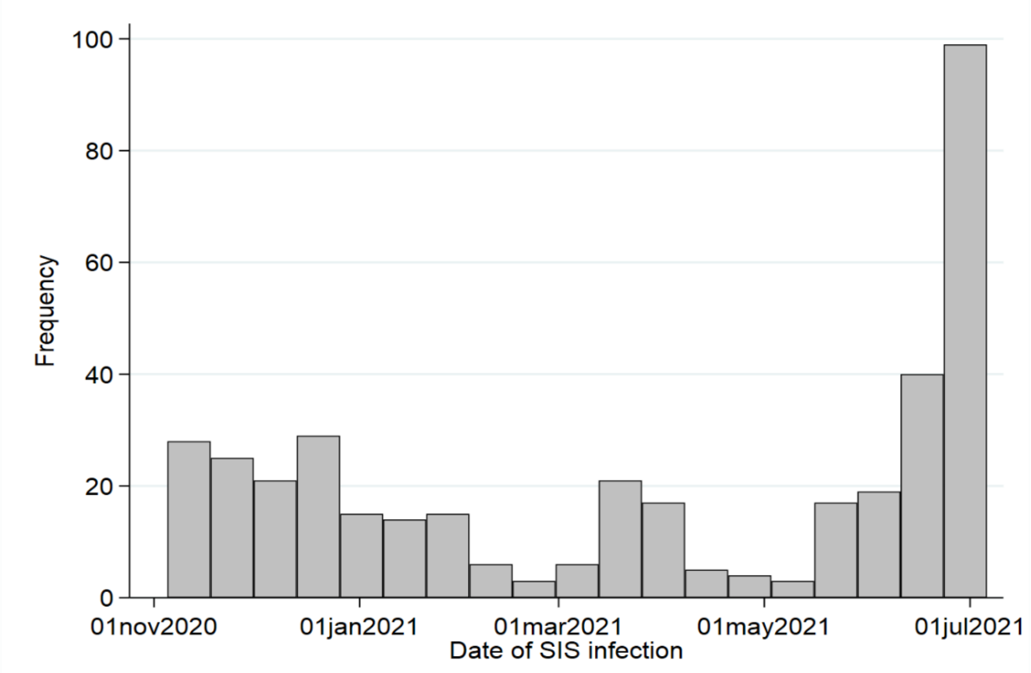


*Supplementary figure S2*: Histogram showing the distribution of laboratory-confirmed SARS-CoV-2 infections amongst staff school infection survey participants in the observation period, Nov 2020 to July 2021 (N=439).

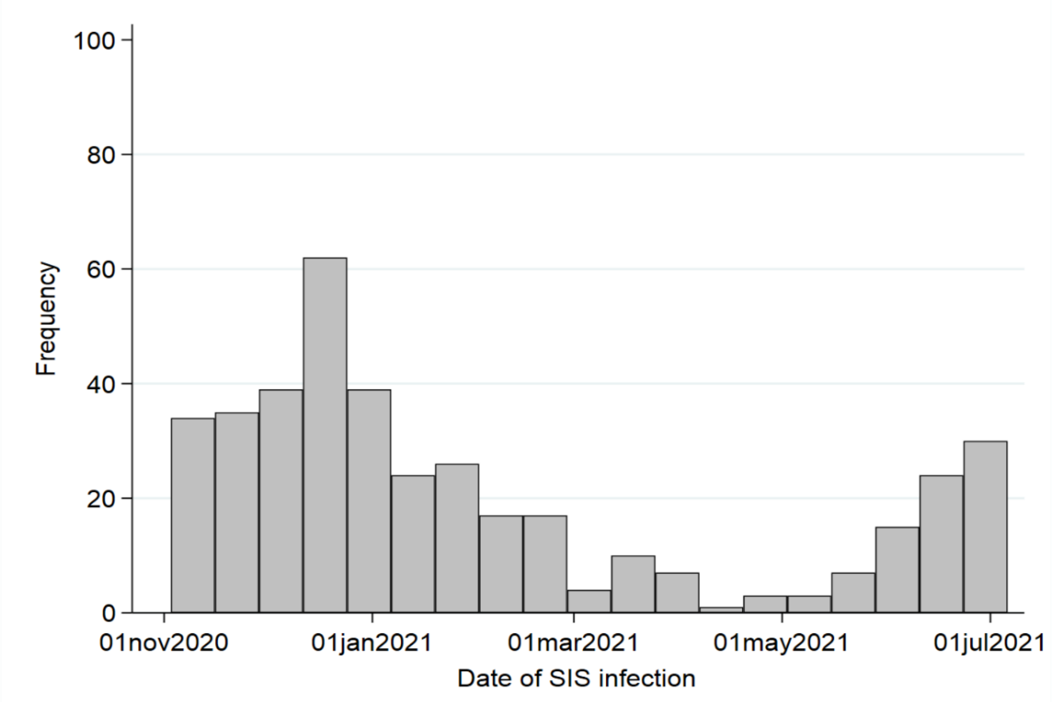

*Supplementary figure S3:* Histograms showing the number of days between participant household infections and school infection survey participant infection for staff and students, Nov 2020 to July 2021.

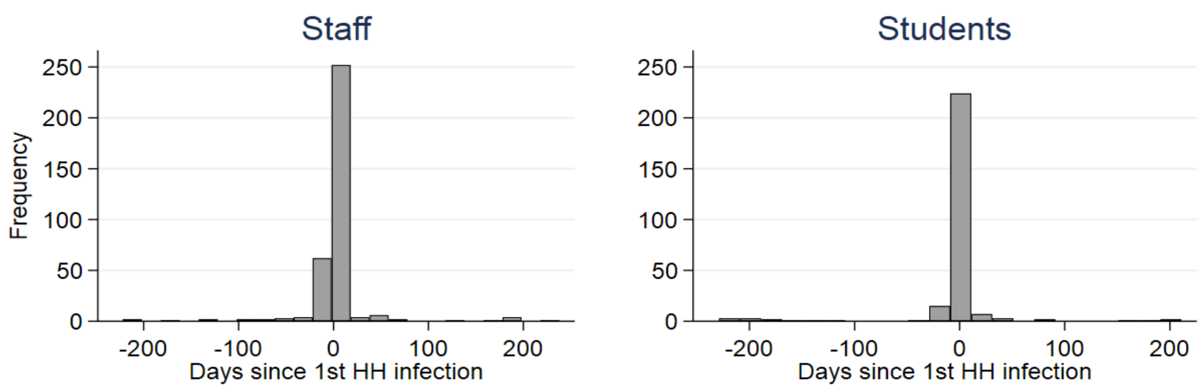


Abbreviations: HH, household; SIS, school infection survey. The graphs show the time in days between SIS participant infections and the first reported household infection (excluding the SIS participant) in the study cohort between 02 November 2020 and 06 July 2021.

### *Supplementary figure S4*: Detail and purpose of the SCCS counterfactual extension used in the main analysis.

A specificity of this study is that SARS-CoV-2 confers some natural immunity, thus censoring study participants' observation period as their risk of re-infection is temporarily and significantly reduced. This potentially violates an important SCCS assumption that the observation period must be independent of event times. To address this, we implemented an extension of the SCCS method in which observation time after SIS participants is replaced by a counterfactual in which no exposure can occur and thus correcting the bias. We implemented the method in Stata by adapting the model to suit the organization of our hypothesized risk periods, and used reweighting and a Poisson pseudo-loglikelihood. Analyses were controlled for time-varying confounding on calendar time and vaccination status (in staff). The SCCS model automatically controls for all observed and unobserved time-invariant confounders.

(Farrington CP, Whitaker HJ, Hocine MN. Case series analysis for censored, perturbed, or curtailed post-event exposures. *Biostatistics*. 2008;10(1):3-16. doi:10.1093/biostatistics/kxn013)

*Supplementary table S1*: Incidence rate ratios (IRR) school infection survey participant infection in relation to a participant household infection, stratified by staff and students, including additional risk periods 13-18 days pre-household infection and 15-21 following household infection, Nov 2020 to July 2021.

| | **Risk group, days surrounding HH infection** | **No of events** | **IRR (95% CI)** | **p-value** |
|---|---|---|---|---|
| **Staff** | Baseline period | 50 | 1 (ref) | - |
| | 13-18 days before | 3 | 2.76 (1.04-7.31) | 0.041 |
| | 6-12 days before | 33 | 18.46 (11.09-30.72) | <0.001 |
| | 0-5 days before | 71 | 70.30 (46.27-106.82) | <0.001 |
| | 1-7 days after | 232 | 111.53 (75.92-163.83) | <0.001 |
| | 8-14 days after | 42 | 14.86 (8.70-25.38) | <0.001 |
| | 15-21 days after | 8 | 4.78 (2.20-10.38) | <0.001 |
| **Students (all ages)** | Baseline period | 58 | 1 (ref) | - |
| | 13-18 days before | <3 | 1.08 (0.24-4.85) | 0.915 |
| | 6-12 days before | 11 | 6.56 (3.19-13.48) | <0.001 |
| | 0-5 days before | 53 | 42.52 (26.53-68.14) | <0.001 |
| | 1-7 days after | 301 | 177.87 (121.49-260.42) | <0.001 |
| | 8-14 days after | 88 | 44.51 (28.16-70.37) | <0.001 |
| | 15-21 days after | 6 | 3.46 (1.29-9.27) | 0.013 |

Abbreviations: CI, confidence interval; HH, household; IRR, incidence rate ratio. IRRs are adjusted for calendar time (all subgroups) and vaccination status (staff only) and estimates are all relative to baseline periods outside of the risk periods. Wald test two-sided p-values.

Comparing IRRs using a wider (39 days) exposure window to our main analysis' 26 days window, showed much lower relative risks in the additional exposure days -18 to -13 and 15 to 21 in both staff (respectively IRR = 2.8, 95%CI 1.0 to 7.3; and IRR = 4.8, 95%CI 2.2 to 10.4) and students (respectively IRR = 1.1, 95%CI 0.2 to 4.9; and IRR = 3.5, 95%CI 1.3 to 9.3); close to baseline and therefore supporting our exposure window definition (*Supplementary table 1*).

*Supplementary table S2*: Incidence rate ratios (IRR) school infection survey participant infection in relation to participant household infection, stratified by staff and students and by potential effect modifiers of interest, using two risk groups amongst subgroups (in the four risk period main analysis, data sparsity did not allow examination of subgroups for age in staff), Nov 2020 to July 2021.

| **Subgroup** | **Potential effect modifier** | **Risk group, days surrounding HH infection** | **No of events** | **Stratum-specific IRR (95% CI)*** | **p-value*** |
|---|---|---|---|---|---|
| Staff | Male sex | Baseline period | 16 | 1 (ref) | - |
| | | 0-12 days before | 19 | 25.85 (12.15-54.99) | <0.001 |
| | | 1-14 days after | 54 | 44.63 (20.76-95.94) | <0.001 |
| | Female sex | Baseline period | 45 | 1 (ref) | - |
| | | 0-12 days before | 85 | 37.49 (24.08-58.38) | <0.001 |
| | | 1-14 days after | 220 | 60.90 (40.23-92.21) | <0.001 |
| | Age <40 | Baseline period | 23 | 1 (ref) | - |
| | | 0-12 days before | 48 | 41.67 (21.98-79.01) | <0.001 |
| | | 1-14 days after | 116 | 68.48 (36.11-129.88) | <0.001 |
| | Age 40+ | Baseline period | 38 | 1 (ref) | - |
| | | 0-12 days before | 55 | 31.31 (19.32-50.75) | <0.001 |
| | | 1-14 days after | 156 | 54.09 (34.36-85.13) | <0.001 |
| | HH size; 1-2 | Baseline period | 12 | 1 (ref) | |
| | | 0-12 days before | 28 | 49.33 (20.52-118.56) | <0.001 |
| | | 1-14 days after | 60 | 71.89 (29.62-174.50) | <0.001 |
| | HH size; 3-4 | Baseline period | 40 | 1 (ref) | |
| | | 0-12 days before | 60 | 29.68 (18.46-47.70) | <0.001 |
| | | 1-14 days after | 155 | 43.89 (28.12-68.50) | <0.001 |
| | HH size; 5+ | Baseline period | 9 | 1 (ref) | |
| | | 0-12 days before | 15 | 34.88 (13.22-92.00) | <0.001 |
| | | 1-14 days after | 59 | 104.05 (40.34-268.36) | <0.001 |
| | Primary school | Baseline period | 22 | 1 (ref) | - |
| | | 0-12 days before | 39 | 39.81 (20.82-76.13) | <0.001 |

| | | | | | |
|---|---|---|---|---|---|
| | | 1-14 days after | 70 | 41.48 (22.44-76.67) | <0.001 |
| | Secondary school | Baseline period | 39 | 1 (ref) | - |
| | | 0-12 days before | 65 | 33.91 (21.05-54.63) | <0.001 |
| | | 1-14 days after | 204 | 72.02 (45.37-114.33) | <0.001 |
| Students | Male sex | Baseline period | 27 | 1 (ref) | - |
| | | 0-12 days before | 35 | 30.29 (14.89-61.58) | <0.001 |
| | | 1-14 days after | 178 | 128.75 (70.16-236.24) | <0.001 |
| | Female sex | Baseline period | 38 | 1 (ref) | - |
| | | 0-12 days before | 29 | 12.28 (6.77-22.29) | <0.001 |
| | | 1-14 days after | 211 | 87.68 (54.57-140.89) | <0.001 |
| | Age <12 | Baseline period | 19 | 1 (ref) | - |
| | | 0-12 days before | 19 | 16.70 (7.47-37.35) | <0.001 |
| | | 1-14 days after | 165 | 147.25 (77.50-279.78) | <0.001 |
| | Age 12-19 | Baseline period | 46 | 1 (ref) | - |
| | | 0-12 days before | 45 | 19.56 (11.43-33.46) | <0.001 |
| | | 1-14 days after | 224 | 83.87 (52.96-132.81) | <0.001 |
| | HH size; 1-2 | Baseline period | 8 | 1 (ref) | |
| | | 0-12 days before | 7 | 15.15 (4.20-54.73) | <0.001 |
| | | 1-14 days after | 16 | 26.39 (8.27-84.22) | <0.001 |
| | HH size; 3-4 | Baseline period | 34 | 1 (ref) | |
| | | 0-12 days before | 37 | 21.82 (12.09-39.36) | <0.001 |
| | | 1-14 days after | 209 | 103.12 (62.57-169.93) | <0.001 |
| | HH size; 5+ | Baseline period | 23 | 1 (ref) | |
| | | 0-12 days before | 19 | 15.04 (6.46-34.98) | <0.001 |
| | | 1-14 days after | 164 | 137.67 (69.29-273.55) | <0.001 |
| | Primary school | Baseline period | 14 | 1 (ref) | - |
| | | 0-12 days before | 9 | 11.19 (3.60-34.78) | <0.001 |

| | | | | | |
|---|---|---|---|---|---|
| | | 1-14 days after | 93 | 123.37 (53.69-283.47) | <0.001 |
| | Secondary school | Baseline period | 51 | 1 (ref) | - |
| | | 0-12 days before | 55 | 20.36 (12.50-33.16) | <0.001 |
| | | 1-14 days after | 296 | 97.18 (64.02-147.51) | <0.001 |

Abbreviations: CI, confidence interval; HH, household; IRR, incidence rate ratio; SIS, school infection survey. No of events refers to the number of SIS infections occurring in that subgroup of patients, in both the baseline and exposure risk periods. IRRs are adjusted for calendar time (all subgroups) and receipt of first dose vaccination (staff only). Wald test two-sided p-values.

*Supplementary table S3*: Incidence rate ratios (IRR) school infection survey participant infection in relation to a participant household infection, stratified by staff, students, and student age groups, restricted to only PCR-confirmed SARS-CoV-2 infections recorded in the SIS panel surveys, Nov 2020 to July 2021.

| | Risk group, days surrounding HH infection | No of events | IRR (95% CI) | p-value |
|---|---|---|---|---|
| **Staff** | Baseline | 18 | 1 (ref) | - |
| | 0-12 days before | 11 | 4.74 (2.00-11.21) | <0.001 |
| | 1-14 days after | 3 | 0.87 (0.18-4.22) | 0.864 |
| **Student (all ages)** | Baseline | 30 | 1 (ref) | - |
| | 0-12 days before | 16 | 6.05 (2.89-12.67) | <0.001 |
| | 1-14 days after | 3 | 0.13 (0.02-1.08) | 0.059 |
| **Student (<12 years)** | Baseline | 15 | 1 (ref) | - |
| | 0-12 days before | 8 | 5.17 (1.76-15.25) | 0.003 |
| | 1-14 days after | 0 | 0.00 (0.00-.) | 0.997 |
| **Student (12-19 years)** | Baseline | 15 | 1 (ref) | - |
| | 0-12 days before | 8 | 6.91 (2.48-19.25) | <0.001 |
| | 1-14 days after | 3 | 0.22 (0.02-2.21) | 0.198 |

Abbreviations: CI, confidence interval; HH, household; IRR, incidence rate ratio; PCR, polymerase chain reaction; SIS, school infection survey. No of events refers to number of SIS infections occurring across the baseline and risk periods. IRRs are adjusted for calendar time (all subgroups) and vaccination status (staff only). Wald test two-sided p-values.

In the subgroup analyses restricted to SIS participants with PCR-confirmed infection during the School Infection Survey panel surveys, there were 32 eligible staff and 49 students; these people were assumed pre- or post-symptomatic or asymptomatic when present on school premises during the survey visits. Overall, the results suggested higher relative incidence in the 0-12 days before the household member infection, compared to baseline observation periods, for both staff and students, (day -12 to day 0 IRR = 4.74 (95%CI 2.00-11.21) in staff and IRR = 6.05 (95%CI 2.89-12.67) in students, although the magnitude of association was much smaller than in the main analysis (*Supplementary table 3*).

Supplementary table S4: Incidence rate ratios (IRR) school infection survey participant infection in relation to a participant household infection, stratified by staff, students, and student age groups, using the standard self-controlled case-series analysis (ignoring the limiting assumption that observation period should not depend on events), Nov 2020 to July 2021.

| | **Risk group, days surrounding HH infection** | **No of events** | **IRR (95% CI)** | **p-value** |
|---|---|---|---|---|
| **Staff** | Baseline period | 61 | 1 (ref) | - |
| | 6-12 days before | 33 | 17.55 (11.51-26.78) | <0.001 |
| | 0-5 days before | 71 | 65.58 (46.89-91.72) | <0.001 |
| | 1-7 days after | 232 | 102.97 (75.89-139.73) | <0.001 |
| | 8-14 days after | 42 | 15.46 (9.79-24.41) | <0.001 |
| **Student (all ages)** | Baseline period | 65 | 1 (ref) | - |
| | 6-12 days before | 11 | 9.47 (5.22-17.16) | <0.001 |
| | 0-5 days before | 53 | 59.66 (41.02-86.78) | <0.001 |
| | 1-7 days after | 301 | 227.47 (167.74-308.48) | <0.001 |
| | 8-14 days after | 88 | 54.18 (37.13-79.06) | <0.001 |
| **Student (<12 years)** | Baseline period | 19 | 1 (ref) | - |
| | 6-12 days before | 3 | 8.46 (2.83-25.32) | <0.001 |
| | 0-5 days before | 16 | 47.49 (23.73-95.02) | <0.001 |
| | 1-7 days after | 124 | 286.51 (170.04-482.74) | <0.001 |
| | 8-14 days after | 41 | 79.98 (43.57-146.83) | <0.001 |
| **Student (12-19 years)** | Baseline period | 46 | 1 (ref) | - |
| | 6-12 days before | 8 | 9.97 (4.90-20.27) | <0.001 |
| | 0-5 days before | 37 | 65.58 (41.87-102.73) | <0.001 |
| | 1-7 days after | 177 | 198.45 (135.96-289.66) | <0.001 |
| | 8-14 days after | 47 | 41.27 (25.15-67.73) | <0.001 |

Abbreviations: CI, confidence interval; HH, household; IRR, incidence rate ratio; SIS, school infection survey. No of events refers to number of SIS participant infections occurring across the baseline and risk periods. IRRs are adjusted for calendar time (all subgroups) and vaccination status (staff only). Wald test two-sided p-values.
